# Association Between Seasonal Respiratory Virus Activity and Invasive Pneumococcal Disease in Central Ontario, Canada

**DOI:** 10.1101/2024.09.03.24312990

**Authors:** Alison E. Simmons, Isha Berry, Sarah A. Buchan, Ashleigh R. Tuite, David N. Fisman

**Affiliations:** Dalla Lana School of Public Health, University of Toronto, Toronto, Ontario, Canada; Centre for Immunization Surveillance and Programs, Public Health Agency of Canada, Ottawa, Ontario, Canada; Health Protection, Public Health Ontario, Toronto, Ontario, Canada

## Abstract

**Background:** In central Ontario, influenza, respiratory syncytial virus (RSV), and invasive pneumococcal disease (IPD) follow similar seasonal patterns, peaking in winter. We aimed to quantify the independent and joint impact of influenza A, influenza B, and RSV on IPD risk at the population level.

**Methods:** We used a 2:1 self-matched case-crossover study design to evaluate acute effects of respiratory virus activity on IPD risk. This design ensures that effects are not confounded by within-individual characteristics that remain constant over short periods of time. We included 3,892 IPD cases occurring between January 2000 and June 2009. Effects were measured using univariable and multivariable conditional logistic regression. Multivariable models included environmental covariates (e.g., temperature, absolute humidity, and UV index) and interaction terms between viruses.

**Results:** Influenza A activity and influenza B activity were both independently associated with increased IPD risk; however, co-circulation of influenza A and B reduced the impact of both viruses. RSV activity was positively associated with increased IPD risk only in the presence of increased influenza A or influenza B activity.

**Conclusions:** To our knowledge this represents the first study to consider the impact of interactions between these viruses on IPD risk in Canada. Our findings suggest that the prevention of IPD should be considered as a potential health benefit of influenza and RSV vaccination programs.

## INTRODUCTION

Invasive pneumococcal disease (IPD), caused by the bacterium *Streptococcus pneumoniae*, is a leading cause of morbidity and mortality among children, older adults, and those with medical comorbidities (1, 2). Between 20 and 60% of children are asymptomatically colonized with *S. pneumoniae* (3), with colonization tending to peak in winter (4). *S. pneumoniae* is opportunistic and, in rare cases, the bacterium invades a normally sterile site causing invasive disease such as bacteremic pneumonia, septicemia, and meningitis.

An association between seasonal respiratory pathogens, including influenza and respiratory syncytial virus (RSV), and IPD has been identified in many global regions including Canada (5-10). Multidirectional relationships between influenza, RSV, and IPD have been observed during the H1N1 influenza pandemic and the COVID-19 pandemic. A decrease in RSV activity following the 2009 H1N1 influenza pandemic at the population level was attributed to viral interference; infection with H1N1 is hypothesized to decrease an individuals’ susceptibility to RSV, shorten the length of the RSV infectious period when co-infected, or reduce their risk of spreading RSV when co-infected (11). In France (12) and Israel (13) declines in influenza and RSV circulation early in the COVID-19 pandemic were associated with declines in IPD risk, despite persistent pneumococcal carriage. By contrast, a 2021 RSV outbreak in Québec, Canada was associated with an increase in IPD among children (8). However, these observations have limited utility in assessment of causality: non-pharmaceutical interventions and decreased population mobility during pandemics may themselves have influenced the epidemiology of multiple respiratory pathogens. Outside the context of pandemics, seasonal co-occurrence of multiple respiratory infections makes it challenging to differentiate correlations from causal relationships.

To overcome these limitations, we previously made use of self-matched epidemiological designs, notably the case-crossover design (6, 7, 14, 15) to evaluate the impact of influenza on IPD risk in a manner that is not confounded by seasonality, propensity for diagnostic testing, and patient-level epidemiological factors such as age, sex, or comorbidity. We found the seasonality of IPD to exhibit consistent seasonal oscillation, in contrast to more irregular seasonal influenza waves in central Ontario (6). However, weekly surges in influenza activity were associated with surges in IPD risk in the following week (6). We subsequently found this association to be generalizable across countries and continents (7). Linkage between influenza-like illness and IPD risk has also been identified by Domenech de Cellès and colleagues using a dynamic transmission model approach (16). We previously used a case-crossover design to study invasive meningococcal disease (IMD) and found that both influenza virus activity and RSV activity were associated with increased IMD risk (14). However, we had not previously evaluated associations between RSV and IPD risk. Furthermore, we previously evaluated pooled influenza activity in a manner that did not distinguish, or consider the possibility of heterogeneity in effects, between influenza A and influenza B viruses.

To fill these gaps, we re-analyzed the central Ontario IPD series from our earlier work, evaluating the effects of influenza A and B viruses separately, and also evaluating the effects of RSV exposure. We accounted for the influence of environmental exposures in our analysis; the relationship between these respiratory viruses and IPD may be confounded by shared seasonality. We hypothesized that influenza A, influenza B, and RSV would be independently associated with elevated IPD risk. We evaluated possible virus-virus interactions using multiplicative interaction terms. During the period under study (2000 to 2009), 7-valent pneumococcal conjugate vaccines (PCV-7) were recommended in Canada for use in pediatric populations (17) and funded in the province of Ontario beginning in January 2005 (18). We evaluated possible changes in the strength of associations between respiratory virus activity and IPD with the introduction of this vaccine.

## METHODS

### Data

Invasive pneumococcal disease cases were obtained from a dataset that has been described and analyzed previously (6, 7). Briefly, IPD cases occurring in the Toronto and Peel regions of central Ontario reported to provincial public health agencies between 2000 and 2009 were used for analysis. Over 30% of the Ontario population lived within these health regions in 2006 (19). The case definition for IPD was clinical evidence of invasive disease (e.g., pneumonia with bacteremia, bacteremia without a known site of infection, meningitis) and the isolation of *S. pneumoniae* or identification of *S. pneumoniae* DNA from a normally sterile site (20). Our analytic dataset contained only anonymized case dates and no individual-level covariates (e.g., age, sex, ethnicity, comorbidity, or clinical outcomes).

Provincial weekly influenza A, influenza B, and RSV test volumes and results were obtained from FluWatch, a sentinel laboratory-based surveillance system administered by the Public Health Agency of Canada’s Respiratory Virus Detection Surveillance System (21). Weekly viral activity levels were applied to all days included in the epidemiological week in which those data were reported. Daily meteorological exposure data for Toronto and Peel were obtained from Environment Canada (22), which reports daily weather conditions from the Toronto Pearson Airport which sits on the border between Peel and Toronto regions. Data on ambient ultraviolent (UV) radiation, reported as a UV index, were obtained from the World Ozone and Ultraviolet Radiation Data Centre (23). Relative humidity measures were converted to measures of absolute humidity (*g*/*m*^3^) using temperature and relative humidity. Environmental covariates incorporated into multivariable models included daily mean temperature, mean absolute humidity, and maximum UV index.

### Design and Analysis

The co-seasonality of IPD and respiratory viruses makes causal inference challenging; correlations between viral and bacterial disease risk and seasonally varying environmental exposures are expected. We examined the seasonality of IPD, respiratory virus activity, and environmental exposures graphically and using fast Fourier transforms (24, 25). To make inferences about the association between respiratory virus activity and IPD, we used a 2:1 matched case-crossover design (6, 7, 14, 15). This approach compares the day on which an IPD case occurred (“case day”) to the other matched days-of-the-week within a 21-day time stratum on which that case did not occur (“control days”). The self-matching implicit in the case-crossover design means that effects are not confounded by within-individual and population characteristics that remain constant over short periods of time (26). Time-matching ensures that effects are not confounded by factors such as non-specific seasonality (7). The random directionality implicit in this approach (control days can follow, precede, or straddle case days) removes biases due to temporal trends in exposures (27). A diagram exemplifying control selection is displayed in Supplementary Figure 1.

Our earlier work suggested that IPD risk was associated with respiratory virus activity in the week prior to IPD case occurrence (6). As the incubation period for pneumococcal disease is thought to be one to three days, we used a hazard period (person-time during which a potential exposure occurred) (26) of one to three days prior to case occurrence. Each stratum (consisting of a case day and two control days) included in our analysis had at least one measurement for each of the exposures within the hazard period.

We created separate conditional logistic regression models including each viral exposure (influenza A, influenza B, or RSV activity) individually, modeled as the mean weekly viral count across the three days. We divided counts by 100 to ensure exponentiated model coefficients were interpretable (i.e., as the relative odds of IPD per 100 additional weekly viral isolations). We re-ran these models incorporating mean environmental exposures (temperature, absolute humidity, and UV index) with the same hazard period to evaluate the possibility of confounding by environmental conditions.

We also evaluated evidence of virus-virus interactions through creation of multiplicative interaction terms (i.e., influenza A activity∗ RSV activity, influenza B activity∗ RSV activity, influenza A activity∗ influenza B activity, and influenza A activity∗ influenza B activity∗ RSV activity). Our first model included only two-way interactions, and our second model included both two-way and three-way interactions between respiratory viruses.

As conjugate vaccines prevent pneumococcal carriage, we hypothesized that viral effects on IPD risk may be attenuated by the adoption of PCV-7. We evaluated this hypothesis by creating a multiplicative interaction term for each viral exposure, with a value of zero prior to 2005 (i.e., date the publicly funded PCV-7 program began), and one from 2005 onwards.

Last, we conducted sensitivity analyses by running our model with all three viral exposures, two-way interaction terms between them, and environmental covariates using two different hazard periods: eight to ten days and one to seven days. We regarded eight to ten days as implausibly long for short term, transient impacts on IPD epidemiology, and used the analysis to assess our use of a hazard period of one to three days. A hazard period of one to seven days enables comparability with our earlier work (6, 7). Analyses were conducted using Stata version 17 (Stata Corp., College Station, Texas) and R Statistical Software Version 4.3.1 (R Core Team, 2023).

### Ethics

We received ethics approval for our study from the Research Ethics Board at the University of Toronto. Our study includes de-identified secondary data and the need for informed consent was waived by the ethics board.

## Results

In Central Ontario, 3,914 cases of IPD were reported between January 2, 2000 and June 7, 2009. Weekly case counts ranged from 1 to 28, with a mean of 8. Pneumococcal cases exhibited marked seasonality (*p* < 0.001 for seasonality from Poisson model with fast Fourier transforms) (**Figure 1**). Influenza A and B, and RSV virus activity also demonstrated profound wintertime seasonality (**Figure 2**). Additionally, environmental covariates displayed marked seasonality (**Supplementary Figure 2**).

**Figure 1.**
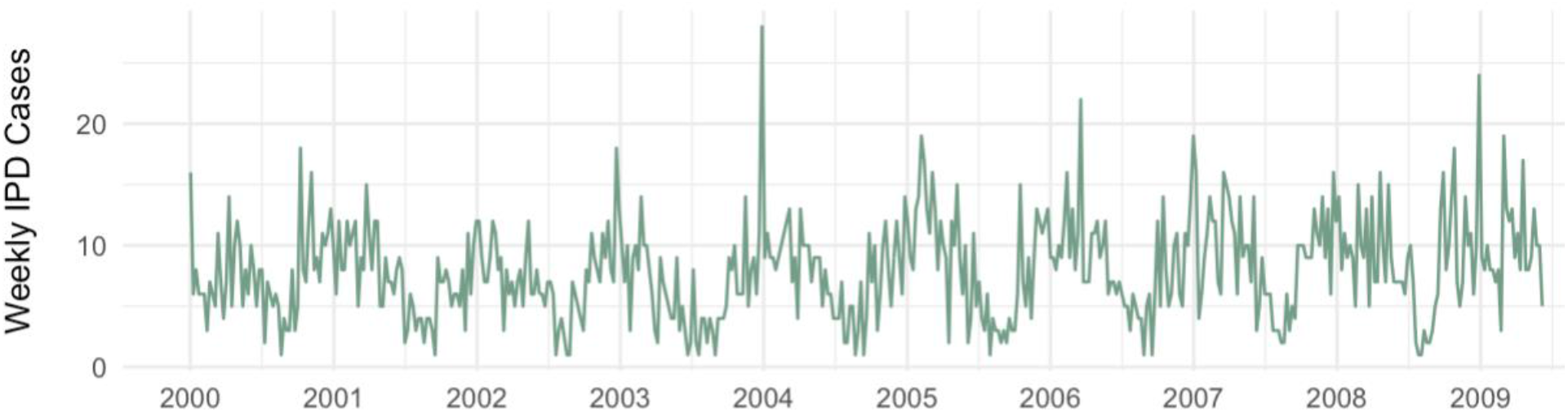
Weekly invasive pneumococcal disease (IPD) cases in Central Ontario, Canada *Notes*: The x-axis labels align with January of each year.

**Figure 2.**
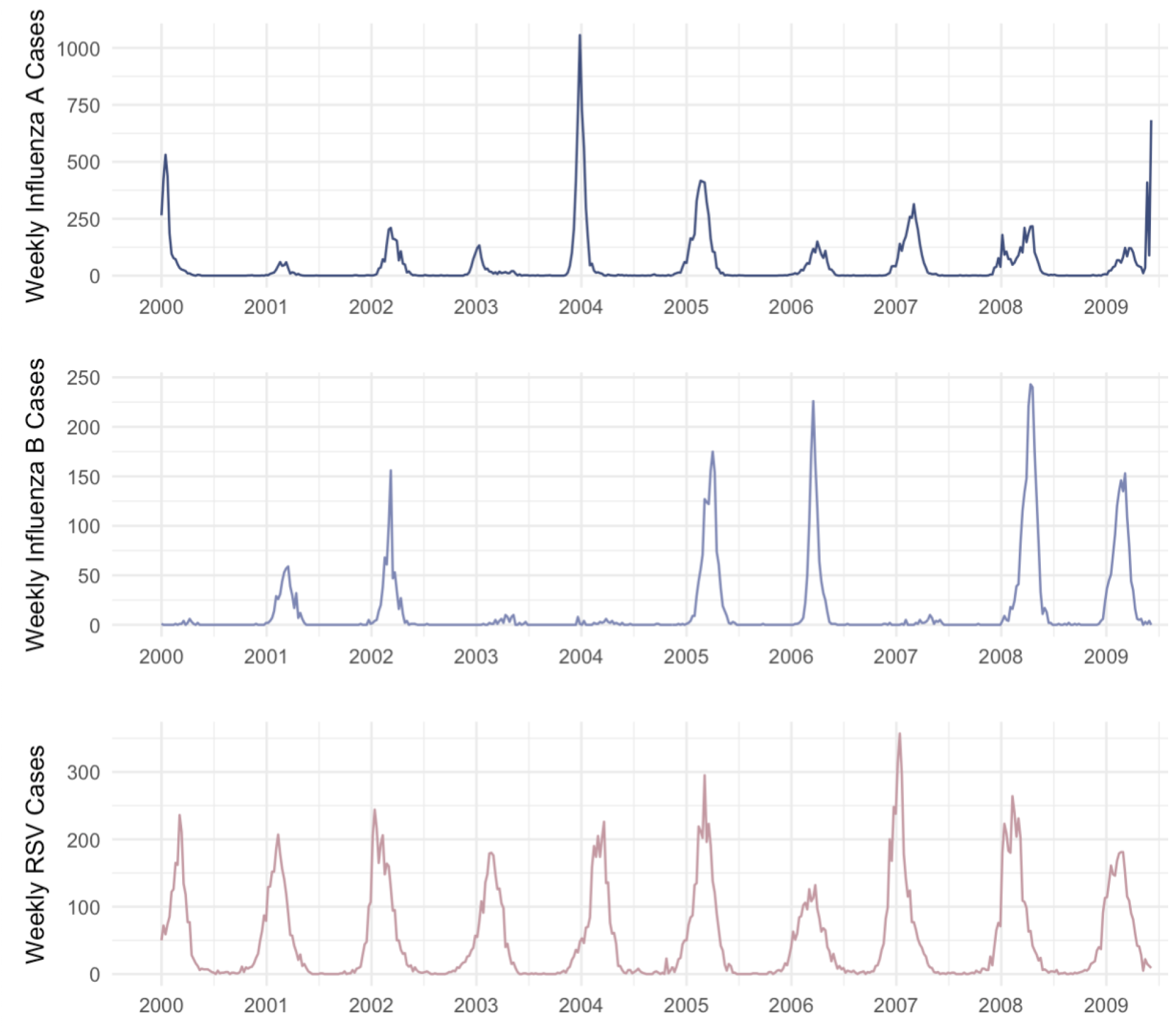
Weekly influenza and respiratory syncytial virus (RSV) cases in Central Ontario, Canada. *Notes*: The x-axis labels align with January of each year. Each graph has a different y-axis scale.

Of the 3,914 reported IPD cases, 3,892 IPD cases were included in our analysis due to missing values of UV index (*n* = 22). The other exposures (i.e., influenza A activity, influenza B activity, RSV activity, absolute humidity, and temperature) had non-missing measures in the hazard period for all cases and controls. In our univariable models, a significant increase in IPD risk was associated with increasing influenza A activity (odds ratio (OR) per 100 isolate increase in weekly viral count = 1.13, 95% CI: 1.04, 1.23) and a trend towards increased IPD risk was seen with increasing influenza B activity (odds ratio (OR) per 100 isolate increase in weekly viral count = 1.24, 95% CI: 0.91, 1.69) (**Table 1**). RSV activity was not associated with IPD risk (OR = 0.94, 95% CI: 0.77, 1.15). Adjustment for UV index, absolute humidity, and mean temperature resulted in no meaningful changes in these estimates. Similarly, in a multivariable model with all viral exposures and environmental covariates modeled simultaneously, no consequential changes were observed.

**Table 1.** Odds ratios (OR) and 95% confidence intervals (95% CI) of the relationship between weekly respiratory virus activity (per 100 viral isolates) and invasive pneumococcal disease (IPD). A hazard period of one to three days between respiratory virus activity and IPD was used.

| Viral exposure | Univariable models |  | Single viral exposure, adjusted for environmental covariates<sup>a</sup> |  | Multivariable model with all viral exposures, adjusted for environmental covariates<sup>a</sup> |  |
| --- | --- | --- | --- | --- | --- | --- |
|  | OR (95% CI) | P-value | OR (95% CI) | P-value | OR (95% CI) | P-value |
| RSV | 0.94<br>(0.77, 1.15) | $p = 0.57$ | 1.00<br>(1.00, 1.00) | $p = 0.55$ | 0.90<br>(0.73, 1.10) | $p = 0.30$ |
| Influenza A | 1.13<br>(1.04, 1.23) | $p < 0.01$ | 1.13<br>(1.04, 1.23) | $p < 0.01$ | 1.13<br>(1.04, 1.23) | $p < 0.01$ |
| Influenza B | 1.24<br>(0.91, 1.69) | $p = 0.17$ | 1.24<br>(0.92, 1.69) | $p = 0.16$ | 1.19<br>(0.88, 1.63) | $p = 0.26$ |

<sup>a</sup> Environmental covariates include mean temperature, mean absolute humidity, and maximum UV index

We explored interactions between viral exposures by incorporating multiplicative interaction terms. Interaction between all three viral exposures were identified (**Table 2**), with non-significant synergistic interaction observed between RSV and influenza A activity, and RSV and influenza B viral activity. Strong antagonism was observed between influenza A and influenza B activity. Influenza A activity and influenza B activity independently increased IPD risk; this risk was further increased in the presence of higher levels of RSV activity. Concomitant high levels of influenza A activity and influenza B activity diminished IPD risk due to antagonism (**Figure 3**). We found no evidence of a three-way interaction between viruses *(p* = 0.32) in a subsequent model that included both two- and three-way interactions, though the negative two-way interaction between influenza A and influenza B activity remained significant (*p* < 0.05) (Not tabled).

**Table 2.** Odds ratios (OR) and 95% confidence intervals (95% CI) of the relationship between weekly respiratory virus activity (per 100 viral isolates) and invasive pneumococcal disease (IPD). A hazard period of one to three days between respiratory virus activity and IPD was used. Absolute humidity, UV index, and temperature were included as covariates.

| Exposure | OR (95% CI) | P-value |
| --- | --- | --- |
| RSV | 0.81 (0.63, 1.05) | $p = 0.11$ |
| Influenza A | 1.11 (0.99, 1.24) | $p = 0.08$ |
| Influenza B | 1.66 (0.89, 3.09) | $p = 0.11$ |
| RSV * Influenza A | 1.08 (0.95, 1.24) | $p = 0.24$ |
| RSV * Influenza B | 1.15 (0.77, 1.70) | $p = 0.50$ |
| Influenza A * Influenza B | 0.74 (0.58, 0.93) | $p < 0.05$ |
| Mean Temperature (°C) | 1.01 (0.99, 1.03) | $p = 0.37$ |
| Mean Absolute Humidity ($g/m^3$) | 0.98 (0.95, 1.02) | $p = 0.39$ |
| Maximum UV Index | 0.98 (0.93, 1.03) | $p = 0.39$ |

**Figure 3.**
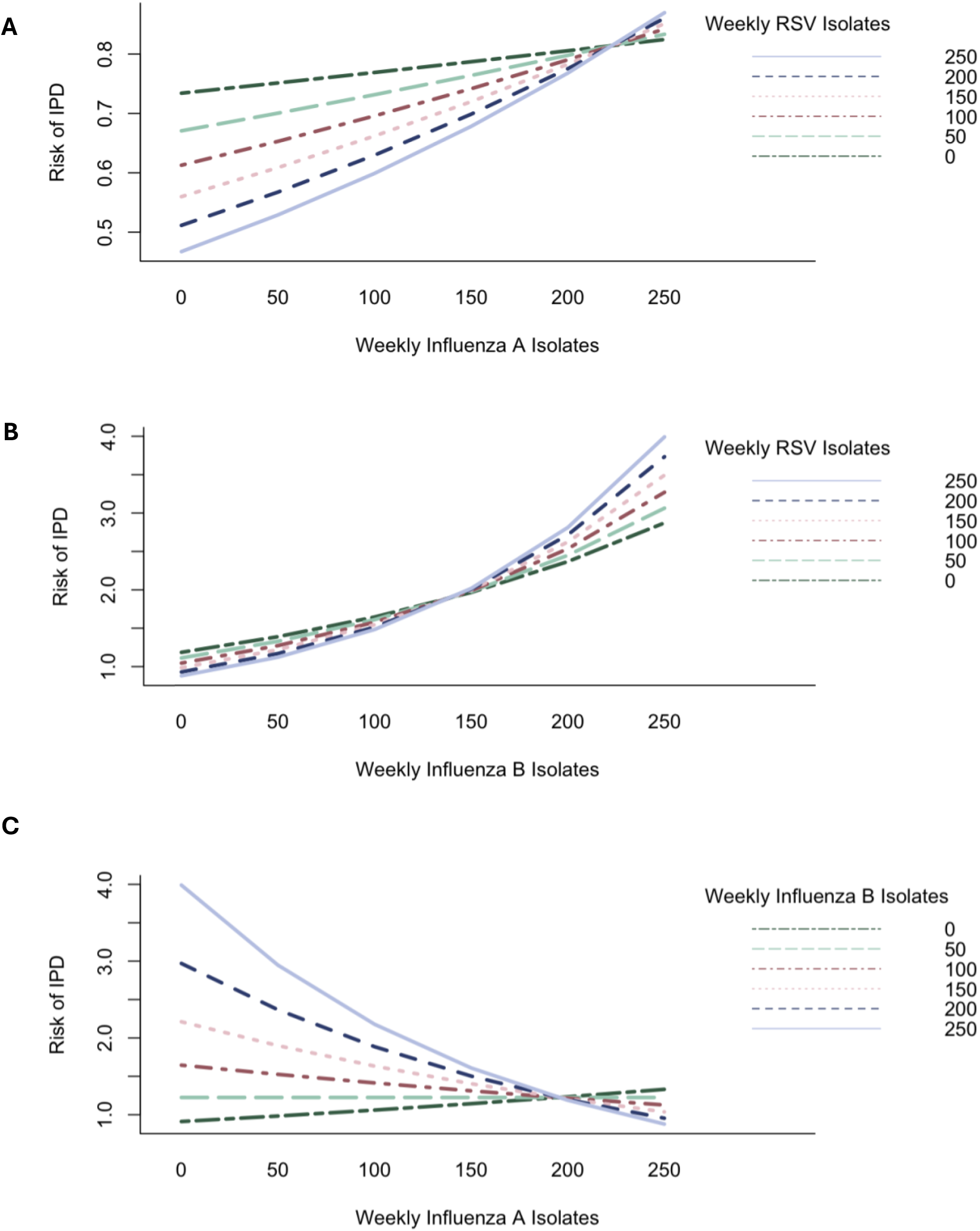
Influence of interactions between weekly isolates of (**A**) influenza A and RSV, (**B**) influenza B and RSV, and (**C**) influenza A and influenza B on IPD risk. There is a positive (synergistic) interaction between influenza A and RSV and influenza B and RSV, and a negative (antagonistic) interaction between influenza A and influenza B.

We hypothesized that the introduction of conjugate pneumococcal vaccination (with a 7-valent pneumococcal conjugate vaccine) may have modified the effect of viral infections on IPD risk. However, we found no significant difference between viral effects on IPD risk in the period from 2000 to 2004 as compared to the period from 2005 to 2009 (RSV activity ∗ PCV-7, *p* = 0.45; influenza A activity ∗ PCV-7, *p* = 0.65; influenza B activity ∗ PCV-7, *p* = 0.31).

In sensitivity analyses using an eight-to-ten-day hazard period to model all three viral exposures and environmental covariates simultaneously, with and without interaction terms, none of the viral exposures were significant (**Supplementary Table 1**). Results from a second sensitivity analysis where a one-to-seven-day hazard period was included, aligning with our team’s earlier work, were not substantially different compared to our primary analysis results (**Supplementary Table 2**).

## DISCUSSION

Building on earlier work, we find that influenza A and influenza B activity significantly affect IPD risk in the Canadian province of Ontario. Our use of a time-matched case-crossover design means that observed effects are not due to seasonal correlation in incidence, nor are they be attributable to characteristics of IPD cases (age, sex, comorbidity) or changing diagnostic practices. To our knowledge this represents the first analysis to consider the effects of influenza A, influenza B, and RSV, and to consider interactions between these viruses on IPD risk. We find these interactions to be complex; increased RSV activity increased IPD risk only in the presence of high influenza A or influenza B activity. Influenza A and influenza B activity appeared to independently increase IPD risk, however, co-circulation of influenza A and B reduced the impact of both viruses on IPD risk. To our knowledge, this represents the first analysis that considers the effects of these three respiratory pathogens on IPD risk in tandem.

Our use of a one-to-three-day hazard (effect) period for action of viral effect represents a refinement of our earlier approach where we used averaged viral activity over the week prior to case occurrence (6, 7). Based on our earlier work, viral effects on IPD risk likely occur as a result of changed susceptibility to invasion, once initial colonization has occurred (6). As confirmed in a sensitivity analysis, this change did not substantially affect our conclusions.

Interaction and interference between viruses is complex and many mechanisms underpin these relationships (28). Infection by one virus may increase or reduce infection risk and replication of a second virus, leading to positive or negative interaction terms in models. The direction and magnitude of interaction between viruses is dependent on the timing of infection with each virus and the hosts’ immune response (28). We identified a positive (synergistic), non-significant interaction between influenza (both A and B) viral activity and RSV viral activity. Prior cellular and epidemiologic studies have shown mixed evidence on the impact of influenza and RSV co-infection on viral replication (28-31), and their impact on risk of bacterial infections (9, 32, 33). We observed a negative (antagonistic), significant interaction between influenza A virus activity and influenza B virus activity. Viral interference and antagonism between influenza A and B has been well described by others (28, 34-36).

We failed to find any modification of respiratory virus effect with the introduction of conjugate pneumococcal vaccines in children in Canada. In retrospect this is perhaps unsurprising given the highest risk group for IPD in Canada is older adults (37, 38), but conjugate pneumococcal vaccines were administered exclusively to young children in Canada during the period under study (17, 18). Although dramatic vaccine-related herd effects during this period resulted in protection of older adults during the period under study (39), these effects would have resulted from prevention of colonization in older adults due to diminished force of infection (due to decreased carriage in children (40)). Consequently, vaccination would not have been expected to modify the effects of respiratory viruses that increase risk of invasion in newly infected individuals, as those protected (indirectly) through herd effects would, by definition, be missing from this analysis (due to lack of colonization). It is possible that effect modification could be seen in vaccinated individuals or in different age groups; however, as our pneumococcal data set consists only of case dates, without information on age, sex or other covariates, we cannot explore that question in this analysis.

In addition to our inability to perform subgroup analyses, other limitations of this analysis include possible non-differential misclassification of exposure. As we are using provincial-level influenza and RSV exposure estimates, but IPD case dates from the province’s major metropolitan area, such that IPD cases were not necessarily exposed to virological or meteorological activity that they have been assigned in our models; this would have the predictable effect of biasing our effects towards the null, so that true effects are likely larger than those that we report here. The fact that we treat viral activity as an ecological exposure might be thought a limitation, but in fact represents a strength of our study design. A large cohort study in which virological testing is performed on a massive scale (such that true individual-level viral infection status upstream from IPD is known) would be complex and resource intensive. More efficient designs, such as those that focus on individuals with both influenza and RSV testing, as well as IPD status, might introduce bias due to correlations between propensity to undergo testing and patient-level characteristics like age, sex and comorbidity. Our use of virological surveillance data as an ecological exposure blocks such paths, as community level surveillance data is not expected to be influenced by an individual’s IPD status. Furthermore, our use of a time-stratified self-matched design means that our results are not confounded by patient-level characteristics, or seasonally varying phenomena (e.g., holidays or the school calendar). We also did not have information on influenza vaccination coverage rates or the annual effectiveness of influenza vaccination. Influenza vaccination may modify the association between influenza and IPD, because vaccinated individuals are further protected from severe illness (i.e., opportunistic infections with *S. pneumoniae)* when infected with influenza (41).

In conclusion, we reaffirmed earlier findings suggesting that viral respiratory infections are highly influential in the genesis of IPD risk. We believe that our finding of complex multi-directional interactions *between* respiratory viruses at a population level, which echo immunological and virological study at the host and cellular level, represents a novel finding. An important gap in this analysis relates to our inability to evaluate modification of these effects by age, sex, or comorbidity. Nonetheless, our findings have important implications for public policy and suggest that prevention of IPD should be considered as a potential health benefit of influenza and RSV vaccination programs.

## Data Availability

Data used in the present study are either publicly available or may be available upon reasonable request from the Toronto Invasive Bacterial Diseases Network.

## SUPPLEMENTARY APPENDIX

**Supplementary Figure 1.**
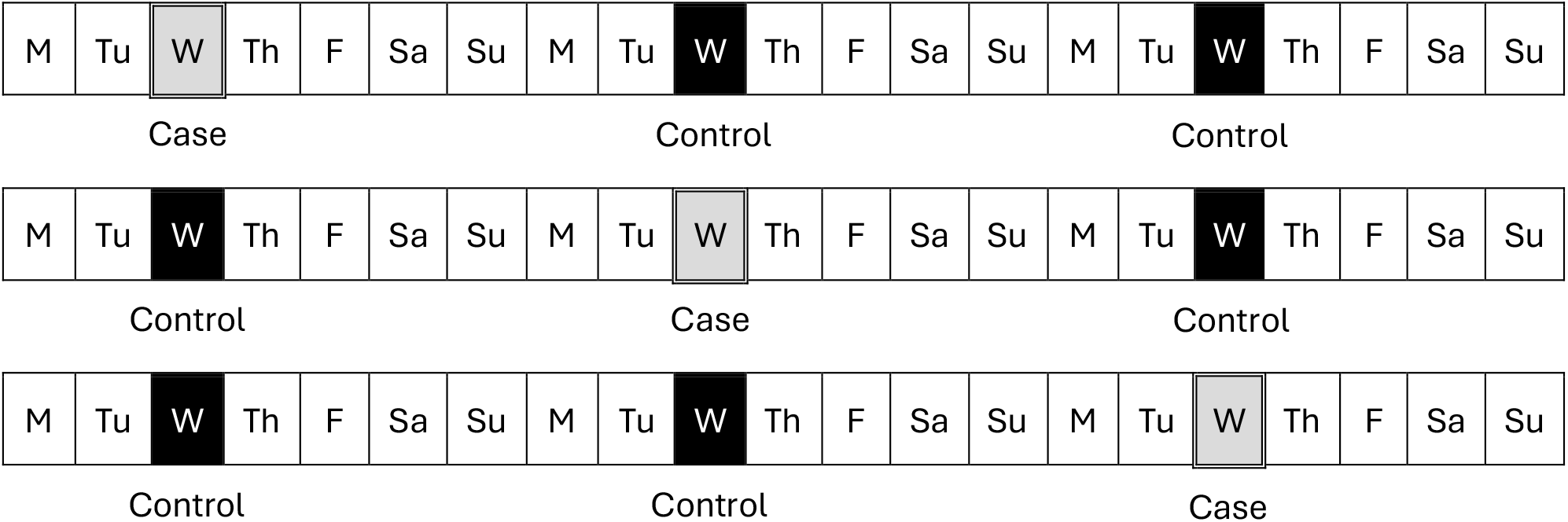
Diagram of control selection for our matched 2:1 case-crossover study, adapted from Berry et al., 2020 (7). Controls were matched with cases by day of the week and could occur up to two weeks before or after a case.

**Supplementary Figure 2.**
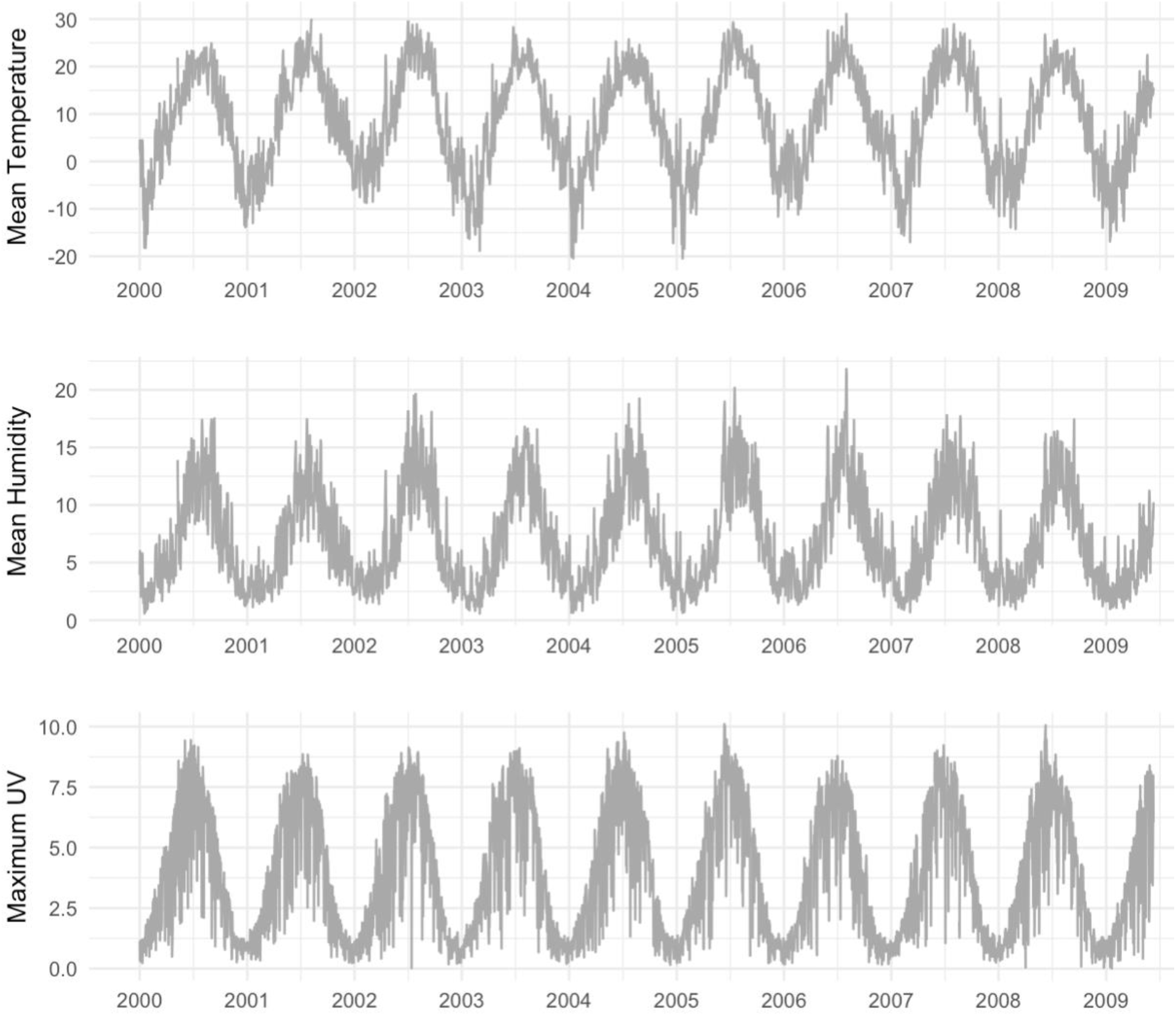
Daily mean temperature (°C), mean absolute humidity (g/m^3^), and maximum UV index in Central Ontario, Canada *Notes*: The x-axis labels align with January of each year. Each graph has a different y-axis scale.

**Supplementary Table 1.**
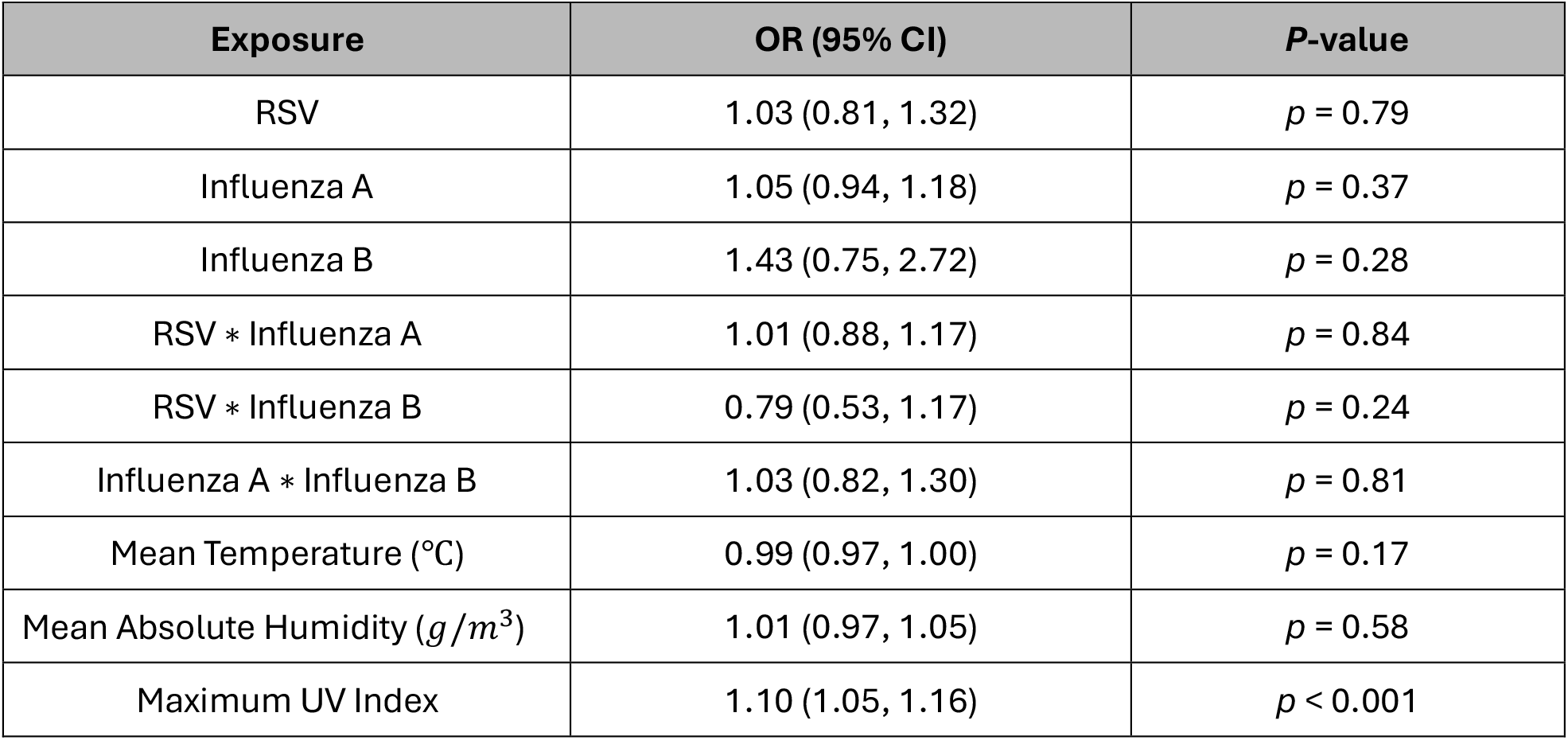
Odds ratios (OR) and 95% confidence intervals (95% CI) of the relationship between weekly respiratory virus activity (per 100 viral isolations) and invasive pneumococcal disease (IPD). A hazard period of eight to ten days between respiratory virus activity and IPD was used. Absolute humidity, UV index, and temperature were included as covariates.

**Supplementary Table 2.** Odds ratios (OR) and 95% confidence intervals (95% CI) of the relationship between weekly respiratory virus activity (per 100 viral isolations) and invasive pneumococcal disease (IPD). A hazard period of one to seven days between respiratory virus activity and IPD was used. Absolute humidity, UV index, and temperature were included as covariates.

| Exposure | OR (95% CI) | P-value |
| --- | --- | --- |
| RSV | 0.75 (0.58, 0.99) | $p < 0.05$ |
| Influenza A | 1.07 (0.95, 1.21) | $p = .26$ |
| Influenza B | 1.65 (0.84, 3.22) | $p = 0.15$ |
| RSV * Influenza A | 1.11 (0.96, 1.29) | $p = 0.16$ |
| RSV * Influenza B | 1.24 (0.81, 1.89) | $p = 0.32$ |
| Influenza A * Influenza B | 0.71 (0.54, 0.92) | $p < 0.05$ |
| Mean Temperature (°C) | 1.01 (0.99, 1.04) | $p = 0.24$ |
| Mean Absolute Humidity ($g/m^3$) | 0.97 (0.92, 1.03) | $p = 0.18$ |
| Maximum UV Index | 0.95 (0.89, 1.02) | $p = 0.31$ |

## Notes

Funding: The doctoral training of AES is supported by an International Doctoral Scholarship from the Connaught Fund at the University of Toronto and a Pandemic Response and Resiliency Award from the Emerging and Pandemic Infections Consortium at the University of Toronto. The funders had no role in study design, data collection and analysis, decision to publish, or preparation of the manuscript.

### Competing Interest Statement

The authors have declared no competing interest.

### Funding Statement

The doctoral training of AES is supported by an International Doctoral Scholarship from the Connaught Fund at the University of Toronto and a Pandemic Response and Resiliency Award from the Emerging and Pandemic Infections Consortium at the University of Toronto.

### Author Declarations

The Research Ethics Board of the University of Toronto gave ethical approval for this work.

## REFERENCES

1. Troeger C, Blacker B, Khalil IA, Rao PC, Cao J, Zimsen SRM, et al. Estimates of the global, regional, and national morbidity, mortality, and aetiologies of lower respiratory infections in 195 countries, 1990–2016: a systematic analysis for the Global Burden of Disease Study 2016. The Lancet Infectious Diseases. 2018;18(11):1191–210.

2. Henriques-Normark B, Tuomanen EI. The pneumococcus: epidemiology, microbiology, and pathogenesis. Cold Spring Harb Perspect Med. 2013;3(7).

3. Public Health Agency of Canada. Invasive Pneumococcal Disease: For Health Professionals 2023 [updated 2023 May 15. Available from: https://www.canada.ca/en/public-health/services/immunization/vaccine-preventable-diseases/invasive-pneumococcal-disease/health-professionals.html.

4. Gray BM, Turner ME, Dillon HC, Jr. Epidemiologic studies of Streptococcus pneumoniae in infants. The effects of season and age on pneumococcal acquisition and carriage in the first 24 months of life. Am J Epidemiol. 1982;116(4):692–703.

5. Weinberger DM, Klugman KP, Steiner CA, Simonsen L, Viboud C. Association between respiratory syncytial virus activity and pneumococcal disease in infants: a time series analysis of US hospitalization data. PLoS Med. 2015;12(1):e1001776.

6. Kuster SP, Tuite AR, Kwong JC, McGeer A, Fisman DN, Network TIBD. Evaluation of coseasonality of influenza and invasive pneumococcal disease: results from prospective surveillance. PLoS medicine. 2011;8(6).

7. Berry I, Tuite AR, Salomon A, Drews S, Harris AD, Hatchette T, et al. Association of Influenza Activity and Environmental Conditions With the Risk of Invasive Pneumococcal Disease. JAMA Network Open. 2020;3(7):e2010167–e.

8. Ouldali N, Deceuninck G, Lefebvre B, Gilca R, Quach C, Brousseau N, et al. Increase of invasive pneumococcal disease in children temporally associated with RSV outbreak in Quebec: a time-series analysis. Lancet Reg Health Am. 2023;19:100448.

9. Li Y, Peterson ME, Campbell H, Nair H. Association of seasonal viral acute respiratory infection with pneumococcal disease: a systematic review of population-based studies. BMJ Open. 2018;8(4):e019743.

10. Weinberger DM, Harboe ZB, Viboud C, Krause TG, Miller M, Mølbak K, et al. Pneumococcal disease seasonality: incidence, severity and the role of influenza activity. European Respiratory Journal. 2014;43(3):833.

11. Li K, Thindwa D, Weinberger DM, Pitzer VE. The role of viral interference in shaping RSV epidemics following the 2009 H1N1 influenza pandemic. medRxiv. 2024:2024.02.25.24303336.

12. Rybak A, Levy C, Angoulvant F, Auvrignon A, Gembara P, Danis K, et al. Association of Nonpharmaceutical Interventions During the COVID-19 Pandemic With Invasive Pneumococcal Disease, Pneumococcal Carriage, and Respiratory Viral Infections Among Children in France. JAMA Netw Open. 2022;5(6):e2218959.

13. Danino D, Ben-Shimol S, van der Beek BA, Givon-Lavi N, Avni YS, Greenberg D, et al. Decline in Pneumococcal Disease in Young Children During the Coronavirus Disease 2019 (COVID-19) Pandemic in Israel Associated With Suppression of Seasonal Respiratory Viruses, Despite Persistent Pneumococcal Carriage: A Prospective Cohort Study. Clinical Infectious Diseases. 2021;75(1):e1154–e64.

14. Tuite AR, Kinlin LM, Kuster SP, Jamieson F, Kwong JC, McGeer A, et al. Respiratory virus infection and risk of invasive meningococcal disease in central Ontario, Canada. PLoS One. 2010;5(11):e15493.

15. Salomon A, Berry I, Tuite AR, Drews S, Hatchette T, Jamieson F, et al. Influenza increases invasive meningococcal disease risk in temperate countries. Clin Microbiol Infect. 2020;26(9):1257 e1–e7.

16. Domenech de Celles M, Arduin H, Levy-Bruhl D, Georges S, Souty C, Guillemot D, et al. Unraveling the seasonal epidemiology of pneumococcus. Proc Natl Acad Sci U S A. 2019;116(5):1802–7.

17. National Advisory Committee on Immunization. Statement on recommended use of pneumococcal conjugate vaccine. Can Commun Dis Rep. 2002;28(Acs-2):1–32.

18. Public Health Ontario. Recommendation for the Routine Pediatric Pneumococcal Immunization Program. 2024. Contract No.: 2024 May 31.

19. Statistics Canada. Table 17-10-0111-01 Estimates of population (2006 Census and administrative data), by age group and sex for July 1st, Canada, provinces, territories, health regions (2013 boundaries) and peer groups.

20. Public Health Agency of Canada. ARCHIVED - Invasive Pneumococcal Disease [updated 2008 May]. Available from: https://www.canada.ca/en/public-health/services/reports-publications/canada-communicable-disease-report-ccdr/monthly-issue/2009-35/definitions-communicable-diseases-national-surveillance/invasive-pneumococcal-disease.html.

21. Public Health Agency of Canada. FluWatch. Ottawa, ON: Public Health Agency of Canada.

22. Environment Canada. Historical Climate Data [updated 2023 Nov 07]. Available from: https://climate.weather.gc.ca.

23. World Ozone and Ultraviolet Radiation Data Centre. [Available from: https://woudc.org/data/explore.php].

24. Scharlemann JP, Benz D, Hay SI, Purse BV, Tatem AJ, Wint GR, et al. Global data for ecology and epidemiology: a novel algorithm for temporal Fourier processing MODIS data. PLoS One. 2008;3(1):e1408.

25. Fisman D. Seasonality of viral infections: mechanisms and unknowns. Clinical Microbiology and Infection. 2012;18(10):946–54.

26. Maclure M. The Case-Crossover Design: A Method for Studying Transient Effects on the Risk of Acute Events. American Journal of Epidemiology. 1991;133(2):144–53.

27. Levy D, Lumley T, Sheppard L, Kaufman J, Checkoway H. Referent Selection in Case-Crossover Analyses of Acute Health Effects of Air Pollution. Epidemiology. 2001;12(2):186–92.

28. Piret J, Boivin G. Viral Interference between Respiratory Viruses. Emerg Infect Dis. 2022;28(2):273–81.

29. Waterlow NR, Flasche S, Minter A, Eggo RM. Competition between RSV and influenza: Limits of modelling inference from surveillance data. Epidemics. 2021;35:100460.

30. Shibata T, Makino A, Ogata R, Nakamura S, Ito T, Nagata K, et al. Respiratory syncytial virus infection exacerbates pneumococcal pneumonia via Gas6/Axl-mediated macrophage polarization. J Clin Invest. 2020;130(6):3021–37.

31. Haney J, Vijayakrishnan S, Streetley J, Dee K, Goldfarb DM, Clarke M, et al. Coinfection by influenza A virus and respiratory syncytial virus produces hybrid virus particles. Nature Microbiology. 2022;7(11):1879–90.

32. Smith CM, Sandrini S, Datta S, Freestone P, Shafeeq S, Radhakrishnan P, et al. Respiratory syncytial virus increases the virulence of Streptococcus pneumoniae by binding to penicillin binding protein 1a. A new paradigm in respiratory infection. Am J Respir Crit Care Med. 2014;190(2):196–207.

33. Martin-Loeches I, van Someren Gréve F, Schultz MJ. Bacterial pneumonia as an influenza complication. Curr Opin Infect Dis. 2017;30(2):201–7.

34. Tobita K, Ohori K. Heterotypic interference between influenza viruses A/Aichi/2/68 and B/Massachusetts/1/71. Acta Virol. 1979;23(3):263–6.

35. Kaverin NV, Varich NL, Sklyanskaya EI, Amvrosieva TV, Petrik J, Vovk TC. Studies on heterotypic interference between influenza A and B viruses: a differential inhibition of the synthesis of viral proteins and RNAs. J Gen Virol. 1983;64 (Pt 10):2139–46.

36. Wanitchang A, Narkpuk J, Jaru-ampornpan P, Jengarn J, Jongkaewwattana A. Inhibition of influenza A virus replication by influenza B virus nucleoprotein: An insight into interference between influenza A and B viruses. Virology. 2012;432(1):194–203.

37. Marrie TJ, Tyrrell GJ, Majumdar SR, Eurich DT. Effect of Age on the Manifestations and Outcomes of Invasive Pneumococcal Disease in Adults. Am J Med. 2018;131(1):100.e1-.e7.

38. Nasreen S, Wang J, Kwong JC, Crowcroft NS, Sadarangani M, Wilson SE, et al. Population-based incidence of invasive pneumococcal disease in children and adults in Ontario and British Columbia, 2002-2018: A Canadian Immunization Research Network (CIRN) study. Vaccine. 2021;39(52):7545–53.

39. Simmons AE, Tuite AR, Buchan S, Fisman D. Pneumococcal Transmission Dynamics During the Use of a Pediatric 13-Valent Pneumococcal Conjugate Vaccine in Canada. SSRN. 2024.

40. O’Brien KL, Millar EV, Zell ER, Bronsdon M, Weatherholtz R, Reid R, et al. Effect of Pneumococcal Conjugate Vaccine on Nasopharyngeal Colonization among Immunized and Unimmunized Children in a Community-Randomized Trial. The Journal of Infectious Diseases. 2007;196(8):1211–20.

41. Pelton SI, Mould-Quevedo JF, Nguyen VH. The Impact of Adjuvanted Influenza Vaccine on Disease Severity in the US: A Stochastic Model. Vaccines (Basel). 2023;11(10).

